# A Deep Transcriptome Meta-Analysis Reveals Sex-based Molecular Differences in Multiple Sclerosis

**DOI:** 10.1101/2021.08.31.21262175

**Authors:** José Francisco Català-Senent, Zoraida Andreu, Marta R. Hidalgo, Francisco José Roig, Natalia Yanguas-Casás, Almudena Neva-Alejo, Adolfo López-Cerdán, Irene Soler-Sáez, María de la Iglesia-Vayá, Francisco García-García

## Abstract

**Background:** Multiple sclerosis (MS), a chronic auto-immune, inflammatory, and degenerative disease of the central nervous system, affects both males and females; however, females suffer from a higher risk of developing MS (2-3:1 ratio compared to males). Current knowledge does not allow a precise definition of the sex-based factors influencing MS. Here, we explore the role of sex in MS to identify potential molecular mechanisms underlying sex-based differences that may guide novel therapeutic approaches tailored for males or females.

**Methods:** We performed a rigorous and systematic review of whole transcriptome studies of MS that included patient information regarding sex in Gene Expression Omnibus and ArrayExpress databases following PRISMA statement guidelines. We analyzed differential gene expression for each selected study and addressed 3 meta-analyses based on genes to evaluate common features and sex bias: the first meta-analysis of 4 nervous tissue studies, a second in 5 blood studies, and a third integrating 9 studies from both tissues. Finally, we performed a gene set analysis on the meta-analyzed differential transcriptomic profiles of the nervous system to study sex-based differences in biological pathways and phenotypes (physiological and pathological states).

**Results:** After screening 122 publications, the systematic review provided a selection of 9 studies (5 in blood and 4 in nervous tissue) with a total of 474 samples (189 MS females and 109 control females; 82 MS males and 94 control males). The tissue-specific meta-analysis identified the overexpression of KIR2DL3 in blood in females and 13 genes with a sex-based differential expression pattern in the nervous system (7 overexpressed in females: ARL17B, CECR7, CEP78, STMP1, TRAF3IP2-AS1, ZNF117 and ZNF488; and 6 overexpressed in males: IFFO2, LOC401127, NUDT18, RNF10, SLC17A5, and UBXN2B). The two-tissue meta-analysis detected a single gene overexpressed in females (LOC102723701). Functional analyses revealed different altered immune scenarios in females and males. A pro-inflammatory environment and innate immune responses related to myeloid linage predominate in females, while in males, adaptative responses associated with the lymphocyte linage. Additionally, MS females displayed alterations in mitochondrial respiratory chain complexes, purine, and glutamate metabolism, while MS males displayed alterations in stress response to metal ion, amine, and amino acid transport.

**Conclusion:** We found transcriptomic and functional differences between MS males and females (especially in the immune system), which may support the development of sex-specific treatments. Our study highlights the importance of understanding sex as a variable in MS.

## Background

Multiple sclerosis (MS), a chronic autoimmune disease of the central nervous system (CNS), is also considered a neurodegenerative disease as progression occurs alongside the degeneration of the CNS, with exacerbated inflammatory reactions causing demyelination. The main symptoms of MS include fatigue, lack of balance, pain, visual and cognitive alterations, speech difficulties, and tremors. MS is mainly diagnosed in young adults (around 20-40 years) and represents the leading cause of disability in this age group that does not derive from trauma [1, 2]. Clinical isolated syndrome (CIS), the earliest diagnosed form of MS, is characterized by 24-hour episodes; meanwhile, relapsing-remitting MS (RRMS) as the most common disease form (85% of people affected) has milder symptoms that tend to appear as brief flare-ups followed by remission for periods of a few weeks or even months. Between 30-50% of patients initially suffering from RRMS develop secondary-progressive MS (SPMS), which is characterized by continuous progression with or without occasional relapses, minor remissions, and phases of stability. Primary progressive MS (PPMS) only affects 10% of all MS patients and is characterized by the absence of definite flare-ups; however, PPMS patients suffer from slow disease onset followed by a steady worsening of symptoms without an intermediate period of remission. Progressive relapsing MS (PRMS), an atypical form, is characterized by clear acute flares, with or without complete recovery, with periods between outbreaks continuously progressing [3]. Finally, 20% of patients suffering from benign MS suffer from mild flare-ups and complete or almost complete remission [4].

While the etiology of MS remains unknown, genetic and environmental factors impact disease-onset, and sex significantly influences many levels [5]. As with other autoimmune and neurodegenerative diseases, MS presents a strong sex bias in incidence, prevalence, symptom severity, disease course, response to therapy, and overall survival [6, 7]. While females have a higher susceptibility to MS than males, males are (on average) older than females at MS diagnosis and suffer from worse and more rapid progression. Males are also more likely to develop progressive MS than females [8, 9]. Sex-based differences in autoimmune and neurological diseases have been previously described [6, 8] and related to the presence of sexual chromosomes (XX vs. XY, gene dosage, imprinting, and transcriptomic profile) [9, 10] and endogenous sex hormones (estrogens, progesterone, androgens, and prolactin) that impact the immune system and promote changes in disease severity. While estrogens represent potent stimulators of autoimmunity, androgens play a protective role [3]. Additionally, the transcriptomic profiles and epigenetic modifications on sex chromosomes influence hormonal actions in immune responses in both sexes. Responses to environmental factors such as microbial exposure, diet, and sociological differences, also represent sex-based factors in MS [11]. Therefore, differences in the prevalence and severity of MS between males and females involve complex and poorly understood interactions between genetic, hormonal, and environmental factors. This knowledge gap, in turn, impairs the development of personalized therapeutics based on sex-specific information.

Multi-omics-based studies in large and clinically well-characterized patient cohorts with accessible sex information data may identify critical pathways and networks related to sexual dimorphism and thereby support the discovery of new biomarkers and targeted sex-specific therapeutic interventions. *In silico* approaches using the large volumes of biological information accessible in public repositories of biomedical data, such as GEO (Genes Omnibus Expression) [12], represent a powerful research tool. In particular, meta-analyses of transcriptomic studies can uncover consensus patterns of expression from several studies focused on the same question due to their capacity for integration and greater statistical power than individual studies.

In this context, we performed a systematic review and selection of transcriptomic studies related to MS from GEO and ArrayExpress databases [12, 13] that included information on patient sex. We analyzed a total of 474 samples from nine different studies (189 MS females, 109 control females, 82 MS males, 94 control males). Given that MS presents neurodegenerative and autoimmune features, the systematic review included studies focused on nervous tissue and blood that reflected the autoimmune profile and could offer a means of discovering non-invasive biomarkers. Comparisons between males and females highlighted common genes involved in MS and differentially-expressed genes related to sex. After differential gene expression (DGE) analysis, we meta-analyzed nervous tissue and blood studies separately and combined in a third meta-analysis. Specific blood and nervous tissue analyses highlighted 1 (KIR2DL3) and 13 (ARL17B, CECR7, CEP78, IFFO2, LOC401127, NUDT18, RNF10, SLC17A5, STMP1, TRAF3IP2-AS1, UBXN2B, ZNF117, ZNF488) differentially-expressed genes that associated with sex in MS patients, respectively. The two-tissue meta-analysis highlighted an RNA gene (LOC102723701) related to sex in MS patients. Subsequent functional enrichment analyses of the nervous tissue data provided vital information about biologic processes in males and females (mainly immune features) that allowed a better understanding of sex-based differences in MS. The identified molecular mechanisms may have important clinical implications for the prevention and treatment of MS and could potentially favor the development of novel, personalized diagnostic and therapeutic applications.

## Methods

### Literature Review and Study Selection

The systematic review was performed by searching for the keyword “multiple sclerosis” on the GEO and ArrayExpress [12, 13] transcriptomic databases. The reviewed period was from January 2002 (generation of the public repositories) to October 2020. Results were filtered by i) organism: “*Homo sapiens*,” ii) study type: “expression profiling by array” or “expression profiling by high throughput sequencing,” and iii) sample count: at least 12 samples. PRISMA Statement guidelines were followed to ensure the quality and transparency of the systematic review [14].

The following exclusion criteria were applied: i) studies not based on MS, ii) experimental design other than patients vs. controls, and iii) the absence of information on patient sex. In addition, studies with patients receiving treatment or who were immunosuppressed were also excluded.

### Bioinformatic Analyses

A defined analytical strategy was followed for each of the selected studies: i) data download and normalization, ii) exploratory analysis, iii) DGE analysis, iv) integration of DGE results using meta-analysis techniques (considering tissue of origin of samples to obtain robust results across all studies), and v) functional enrichment of meta-analyses data (Figure 1). By grouping the studies on a tissue-specific basis or all tissues together, this analysis allowed us to explore sex-based differences in the global disease and blood and nervous tissue. Bioinformatics analysis was carried out using R 4.1.2 [15]; the packages used and their version number are described in Supplementary Table S1 and the following Zenodo repository (http://doi.org/10.5281/zenodo.6344450).

**Figure 1.**
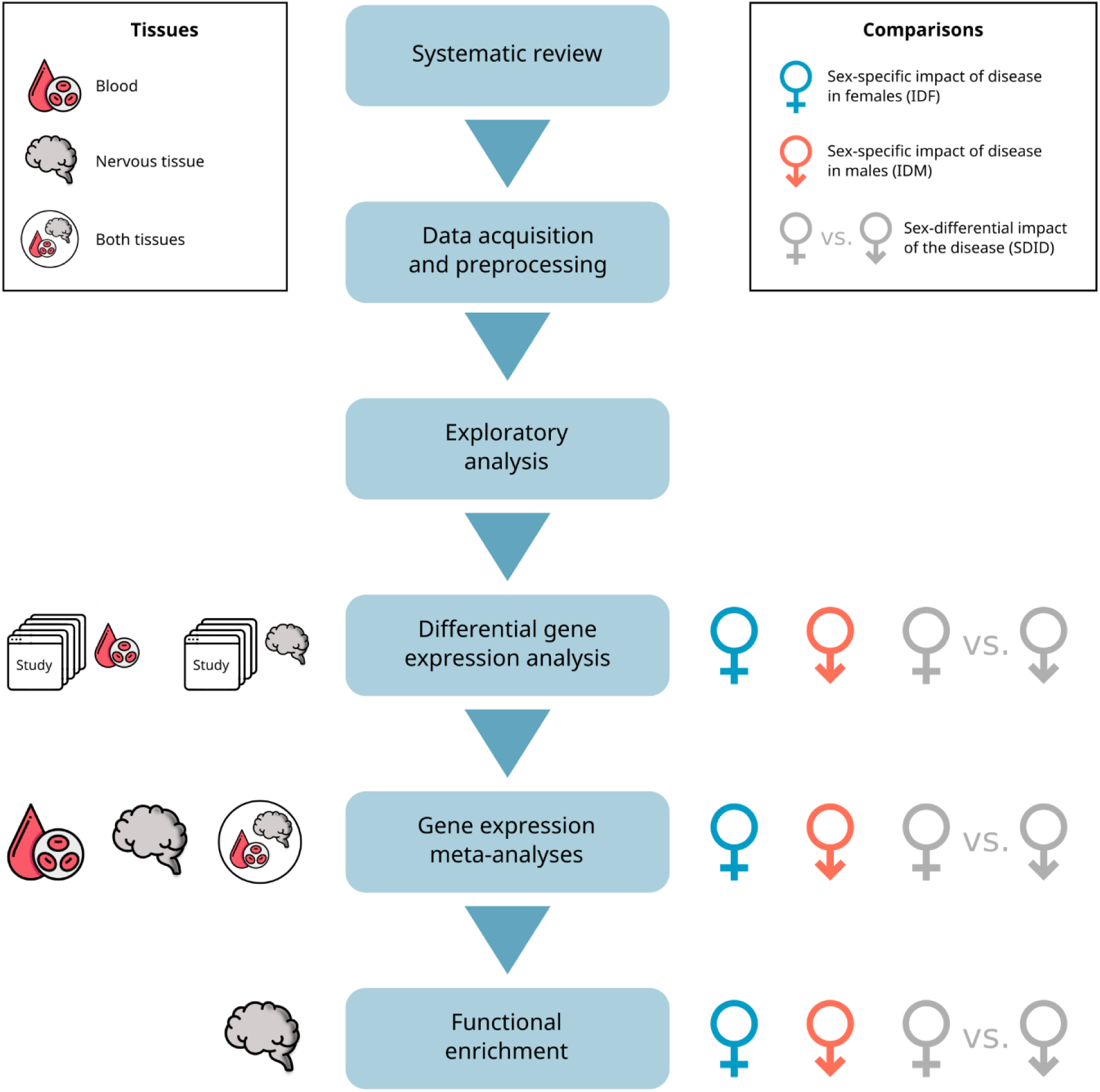
Flowchart describing the work performed, indicating tissues analyzed and comparisons performed in each step.

### Data Acquisition and Preprocessing

First, study data were downloaded and standardized as follows: conversion and updating (if necessary) gene names to HGNC Gene Symbol nomenclature [16], followed by the calculation of the median expression in the case of multiple values for a gene.

The labels used for the sex of the patients were homogenized, and individuals were grouped into healthy or affected by MS. Paired samples were tagged to account for this factor in subsequent analyses. After normalization of the data, possible batch effects or anomalous behavior of the data were analyzed using clustering and principal component analysis (PCA).

### Comparisons

Different comparisons were performed to detect sex-specific genes and find sex-differential functional patterns. The impact of the disease in females (IDF) was analyzed by comparing female MS patients to healthy female controls (Female.MS - Female.Ctrl), while the impact of the disease in males (IDM) was analyzed by comparing males MS patients to healthy male controls (Male.MS - Male.Ctrl). Significant features in both comparisons represent potential disease biomarkers, while those significant in just one comparison can be considered sex-specific biomarkers. Additionally, we analyzed the sex-differential impact of the disease (SDID) through an advanced comparison (Female.MS - Female.Ctrl) - (Male.MS - Male.Ctrl) to find differences in disease impact by sex (Figure 1).

As a measure of the statistical effect, the logarithm of the fold change (LFC) was used for DGE analysis, and the logarithm of the odds ratio (LOR) was used for functional enrichment analysis. Positive statistics indicate that the variable has a higher mean in the first element of the comparison, while negative statistics indicate higher mean values in the second element. Positive LFC or LOR values indicate higher values in MS patients (MS females and MS males in IDF and IDM comparisons) and, therefore, gene upregulation or function overrepresentation in MS. Negative LFC or LOR indicates higher values in control samples (healthy control females and healthy control males), and therefore gene downregulation or function underrepresentation in MS. The SDID comparison, for which each element is a comparison itself, focuses on finding differences between the IDF and IDM comparisons. Thus, a positive statistic could indicate: i) an upregulation in IDF together with a downregulation in IDM, ii) an upregulation in IDF without significant changes in IDM, iii) an upregulation in IDF and IDM but higher in IDF, iv) a downregulation in IDM without a significant change in IDF, or v) a downregulation in IDF and IDM, but higher in IDF (Supplementary Figure 1A-E). A negative statistic associated with the opposite of these cases (Supplementary Figure 1F-J). In this comparison, the particular behavior of each feature across the 4 groups must be assessed *a posteriori*, examining IDF and IDM comparisons. For simplicity, features with positive LFC or LOR will be referred to as features with an incremental change in females or increased in females, and features with a negative LFC or LOR as features with an incremental change in males or increased in males.

### Differential Gene Expression and Meta-analyses

3 DGE analyses were performed for each study to detect differences in gene-level expression between groups following comparisons between IDF, IDM, and SDID as described above. All comparisons were performed using the R package limma [17]. After calculating the differential expression statistics, p-values were adjusted using the Benjamini & Hochberg (BH) method [18].

To robustly integrate the DGE results of each study, the meta-analysis technique was employed for each comparison (IDF, IDM, and SDID) and available tissue of origin (blood, nervous tissue, and both together), which led to a total of 9 meta-analyses. The two-tissue meta-analysis included only genes detected in at least 1 study from each tissue. The meta-analyses were performed following the methodology previously described by García-García [19]. Briefly, the metafor R package [20] combined the expression results of the corresponding individual studies, using a random-effects model for each gene, which is particularly suitable when there exist diverse sources of variability [21]. For the overall calculation of LFC, the least variable results were given greater weight by considering the variability of the individual studies in the global estimation of the measured effect. Better integration of the selected studies and statistically more robust results were achieved by incorporating the variability between experiments in the random-effects model. The suitability of this method was evaluated by assessing the heterogeneity and influence of each study on the overall model and cross-validation techniques.

The meta-analyses calculate a pooled measure of the expression effect across all studies for each gene (combined LFC and its 95% confidence interval) and provide a BH-adjusted p-value [18]. The significance cutoff was set to 0.05. The Open Targets platform [22] (release 21.09.5) was used to evaluate the association of genes with MS.

### Functional Enrichment Analyses

To detect sex-specific and sex-differential functions or pathways associated with MS, a gene set analysis (GSA) [23] of nervous tissue meta-analysis was performed for each of the 3 comparisons (IDF, IDM, and SDID). First, genes were ordered according to the p-value and sign of the statistic obtained in the DGE, and GSA was performed using the logistic regression model implemented in the mdgsa R package [24]. The functional annotation required for GSA was obtained from the following databases: Gene Ontology (GO) [25] in the case of biological processes (BP) and Kyoto Encyclopedia of Genes and Genomes (KEGG) Pathway [26] for pathways. Due to their hierarchical structure, the gene annotations were propagated with GO terms applied in the mdgsa package to inherit the annotations of ancestor terms. Next, excessively specific or generic annotations were filtered out (blocks smaller than 10 or larger than 500 genes). Finally, functions with a Benjamini & Yekutieli (BY) adjusted p-value [27] under 0.05 were considered significant. To obtain a global view of sex-specific alterations in GO biological processes, the GSA results of each comparison were grouped into categories defined by hierarchically superior GO terms, counting the number of processes within each of them.

### Metafun-MS Platform

All data and results generated in the different steps of the meta-analysis are available on the Metafun-MS platform (http://bioinfo.cipf.es/metafun-MS), which is freely accessible to any user and allows the confirmation of the results described in this manuscript and the exploration of other results of interest. The front-end was developed using the Angular Framework. All graphics used in this web resource have been implemented with Plot.ly [28] except for the exploratory analysis cluster plot generated with ggplot2 [29].

This easy-to-use resource is divided into 5 sections: i) the summary of analysis results in each phase, followed by detailed results for the ii) exploratory analysis, iii) DGE, iv) meta-analysis, and v) functional profiling for each study. The user can interact with the web tool through graphics and tables and explore information associated with specific genes or biological functions. In addition, the detailed results of all analyses are available in the public repository Zenodo (http://doi.org/10.5281/zenodo.6344450).

## Results

### Systematic Review and Exploratory Analysis

Our systematic review screened 122 publications, which rendered a total of 9 studies eligible for our analyses: 5 studies with blood samples and 4 studies with nervous tissue samples (Figure 2, Table 1). Notably, we discarded 35% of the studies evaluated for not including sex information or only including individuals of a single sex.

**Figure 2.**
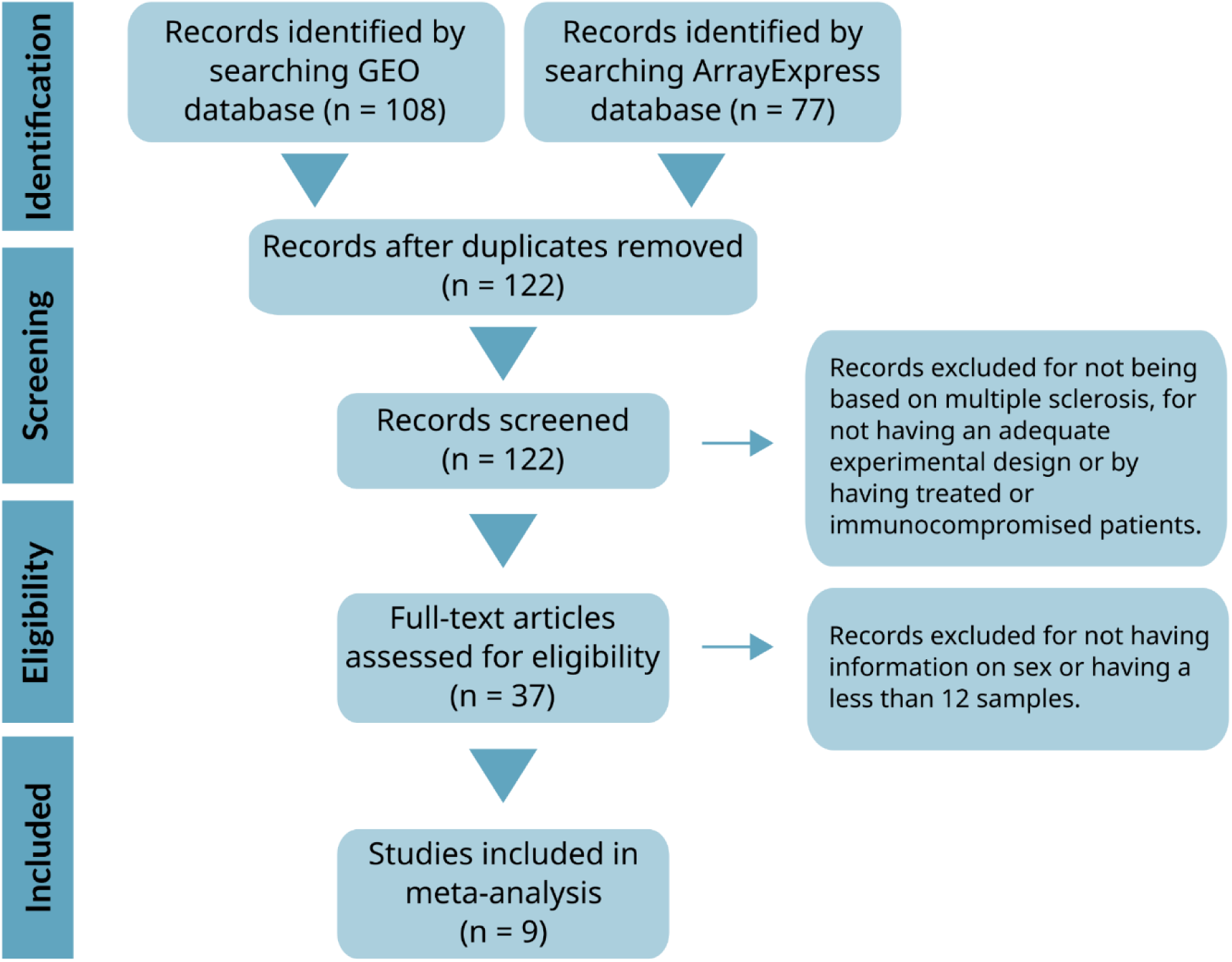
PRISMA diagram of the systematic review.

**Table 1.** Description of selected studies. Platform, tissue, and MS subtype for the samples from 9 studies. The characteristics of the studies are shown as available in the GEO database. An asterisk (MS*) is used when the study does not provide MS subtype information (PPMS - progressive primary multiple sclerosis, SPMS - secondary-progressive multiple sclerosis).

| GEO accession | Platform | Tissue | Subtypes of MS |
| --- | --- | --- | --- |
| GSE37750 [30] | Affymetrix Human Genome U133 Plus 2.0 Array | Plasmacytoid dendritic cells | MS* |
| GSE41848 [31] | Affymetrix Human Exon 1.0 ST Array | Whole blood | Relapse, remission |
| GSE41849 [31] | Affymetrix Human Exon 1.0 ST Array | Whole blood | MS* |
| GSE41890 [32] | Affymetrix Human Gene 1.0 ST Array | Peripheral blood leukocytes | MS* |
| GSE62584 [33] | Affymetrix Human Genome U133 Plus 2.0 Array | CD4+ blood cells, CD8+ blood cells, CD14+ blood cells, and CD19+ blood cells | ON patient in relapse |
| GSE108000 [34] | Agilent-026652 Whole Human Genome Microarray 4x44K v2 | Brain | MS active, MS inactive |
| GSE111972 [35] | Illumina NextSeq 500 | Corpus callosum and occipital cortex | MS* |
| GSE131281 [36] | Illumina HumanHT-12 V4.0 expression beadchip | Grey matter | SPMS, PPMS |
| GSE135511 [37] | Illumina HumanRef-8 v3.0 expression beadchip | Motor cortex | MS lesion, MS normal-appearing |

The 9 selected studies contained 474 samples - 271 MS patients and 203 healthy control patients (62.9% female samples and 37.1% male samples). Table 2 describes the distribution of the samples in detail.

**Table 2.**
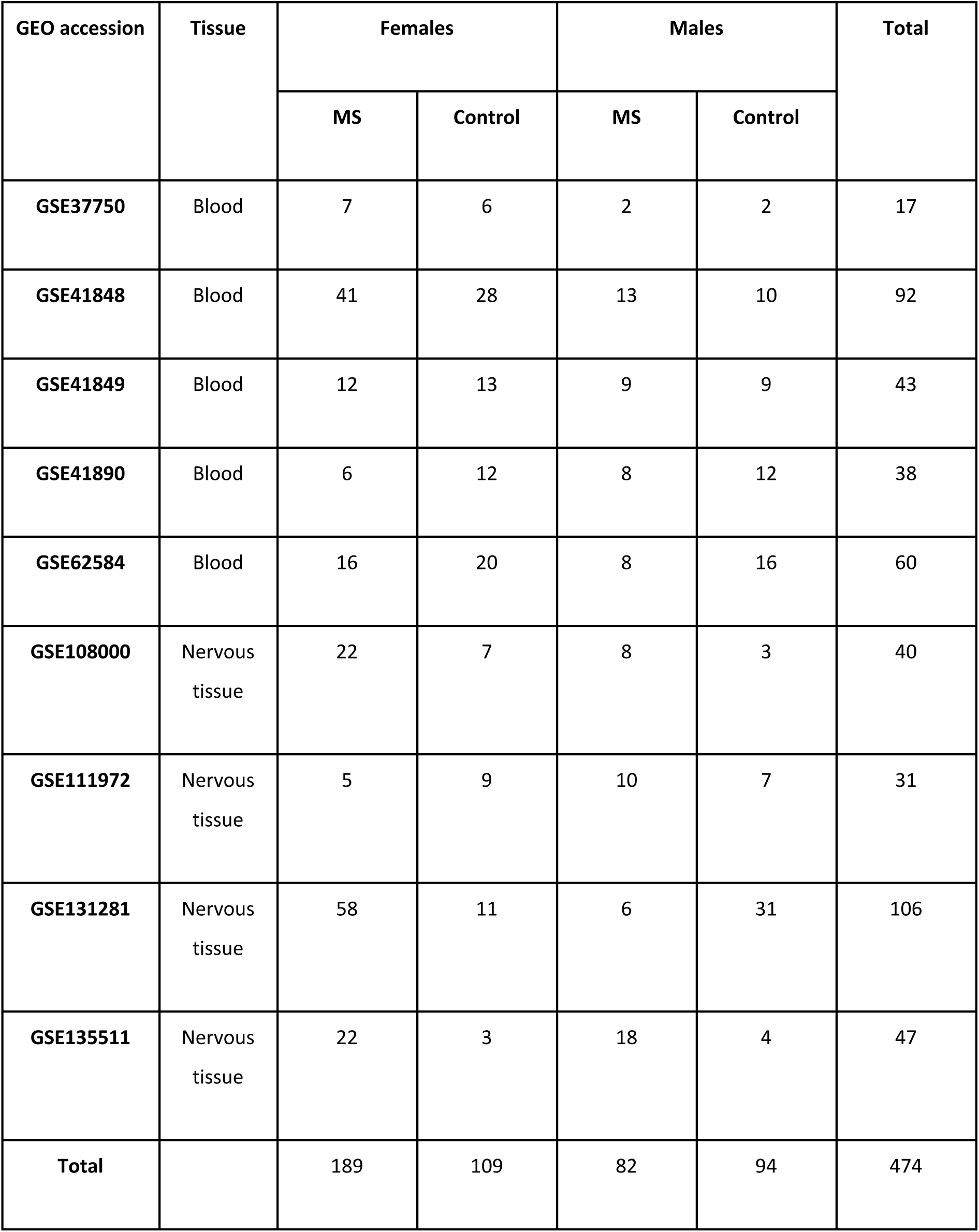
Distribution of samples by sex and experimental group.

| GEO accession | Tissue | Females |  | Males |  | Total |
| --- | --- | --- | --- | --- | --- | --- |
|  |  | MS | Control | MS | Control |  |
| **GSE37750** | Blood | 7 | 6 | 2 | 2 | 17 |
| **GSE41848** | Blood | 41 | 28 | 13 | 10 | 92 |
| **GSE41849** | Blood | 12 | 13 | 9 | 9 | 43 |
| **GSE41890** | Blood | 6 | 12 | 8 | 12 | 38 |
| **GSE62584** | Blood | 16 | 20 | 8 | 16 | 60 |
| **GSE108000** | Nervous tissue | 22 | 7 | 8 | 3 | 40 |
| **GSE111972** | Nervous tissue | 5 | 9 | 10 | 7 | 31 |
| **GSE131281** | Nervous tissue | 58 | 11 | 6 | 31 | 106 |
| **GSE135511** | Nervous tissue | 22 | 3 | 18 | 4 | 47 |
| **Total** |  | 189 | 109 | 82 | 94 | 474 |

Exploratory analyses revealed the absence of biases and anomalous behavior in the samples from all studies except for GSE62584, in which we detected a batch effect produced by the 4 cell types analyzed in that study. We considered this effect during subsequent analyses.

### Differential Gene Expression Analysis

As described in the material and methods section, we evaluated 3 comparisons for each study: IDF, IDM, and SDID. Table 3 summarizes the number of significantly differentially-expressed genes, while the associated Zenodo repository and Metafun-MS web platform contain the detailed results for each comparison.

**Table 3.** The number of significantly differentially-expressed genes by study (and tissue). Description of significant genes by comparisons: IDF (impact of the disease in females), IDM (impact of the disease in males), and SDID (sex-differential impact of the disease). Significant genes are separated according to the sign of their log fold-change (LFC). The final column reports the number of genes assessed in each study.

| Study | Tissue | COMPARISON |  |  |  |  |  | Analyzed genes |
| --- | --- | --- | --- | --- | --- | --- | --- | --- |
|  |  | SDID |  | IDF |  | IDM |  |  |
|  |  | LFC > 0 | LFC < 0 | LFC > 0 | LFC < 0 | LFC > 0 | LFC < 0 |  |
| GSE37750 | Blood | 0 | 0 | 0 | 0 | 0 | 0 | 22,899 |
| GSE41848 | Blood | 0 | 0 | 23 | 9 | 0 | 0 | 17,479 |
| GSE41849 | Blood | 0 | 0 | 0 | 0 | 0 | 0 | 17,479 |
| GSE41890 | Blood | 15 | 15 | 17 | 4 | 93 | 3 | 19,914 |
| GSE62584 | Blood | 145 | 356 | 0 | 0 | 55 | 25 | 13,277 |
| GSE108000 | Nervous | 154 | 230 | 2038 | 3114 | 1811 | 2209 | 21,586 |
| GSE111972 | Nervous | 0 | 0 | 0 | 0 | 0 | 0 | 20,846 |
| **GSE131281** | Nervous | 4 | 0 | 15 | 8 | 0 | 3 | 20,923 |
| **GSE135511** | Nervous | 0 | 0 | 12 | 0 | 100 | 114 | 15,434 |

### Meta-analysis of DGE Profiles

The SDID meta-analysis in the different tissues reported 15 genes that displayed significant alterations between sexes (KIR2DL3 in blood; ARL17B, CECR7, CEP78, IFFO2, LOC401127, NUDT18, RNF10, SLC17A5, STEMP1, TRAF3IP2-AS1, UBXN2B, ZNF117, ZNF488 in nervous tissue; LOC 102723701 in both tissues). We also found that specific genes displayed significance for IDF and/or IDM comparisons, which allowed us to detect the change between sexes and the alterations between healthy and diseased individuals within each sex. We will discuss the alterations found in each tissue in the following sections.

We have made the complete results of the meta-analyses available in the Zenodo repository, the supplementary materials, and on the Metafun-MS web platform. In summary, Figure 3 shows the pattern of LFC change between sexes (SDID, dashed line), in females (IDF, orange), and males (IDM, green). In addition, Table 4 indicates the number of significantly altered genes for each tissue and comparison.

**Figure 3.**
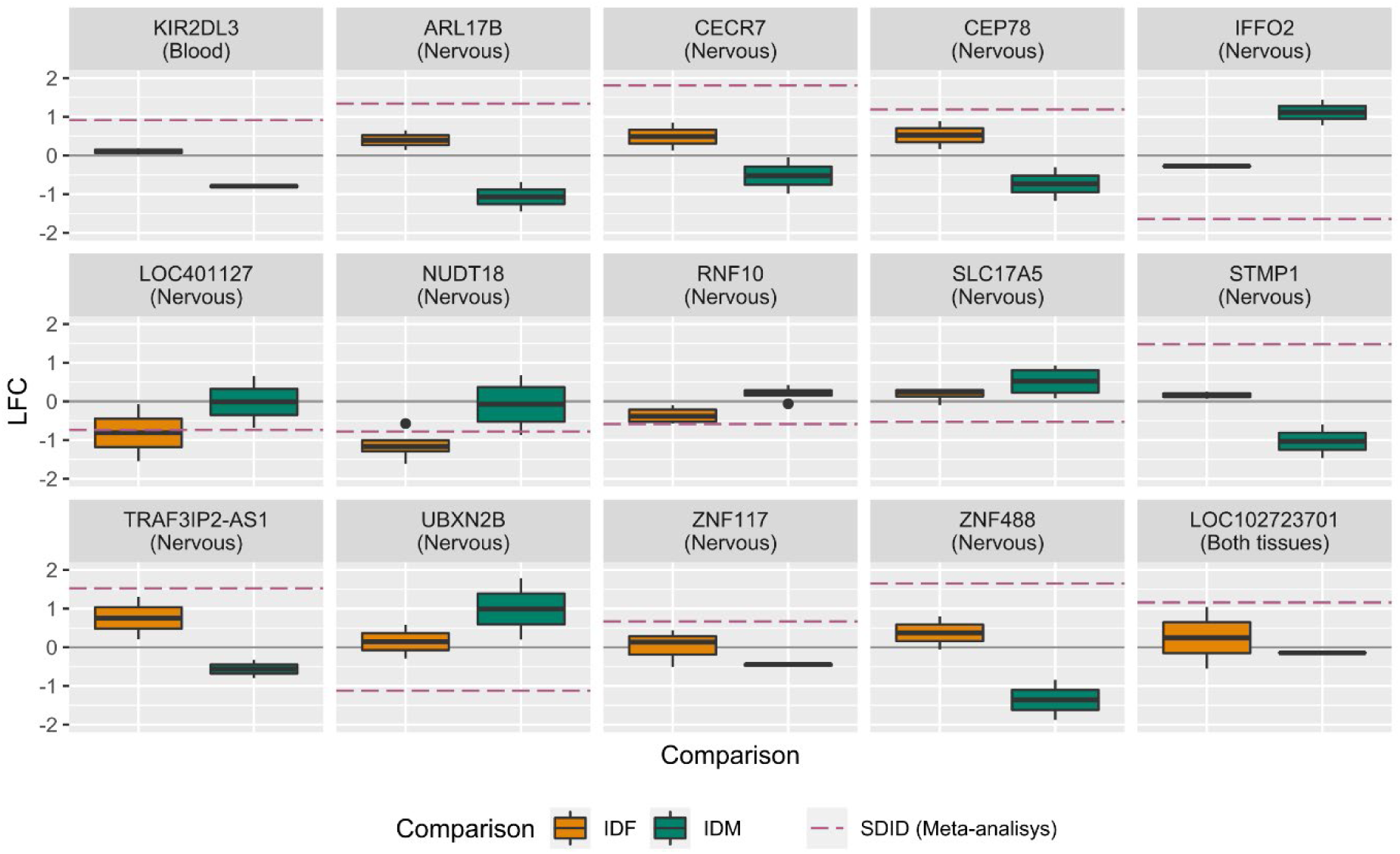
Boxplot of LFC values for the DGE in the SDID comparison. Each boxplot indicates the log fold-change (LFC) values of a gene in the individual DGEs. The dashed purple line indicates the LFC value of the meta-analysis for the SDID comparison. The gray line (at a value of 0) indicates the absence of change. The tissue in which the gene is significant is indicated in parentheses under the gene name.

**Table 4.** Number of DGEs in the meta-analyses by comparison and tissue. Description of significant genes by comparisons: IDF (Impact of the disease in females), IDM (Impact of the disease in males), and SDID (Sex-differential impact of the disease). Significant genes are separated according to the sign of their log fold-change (LFC).

| Tissue | COMPARISON |  |  |  |  |  |
| --- | --- | --- | --- | --- | --- | --- |
|  | SDID |  | IDF |  | IDM |  |
|  | LFC > 0 | LFC < 0 | LFC > 0 | LFC < 0 | LFC > 0 | LFC < 0 |
| **Blood** | 1 | 0 | 5 | 4 | 27 | 5 |
| **Nervous** | 7 | 6 | 33 | 57 | 29 | 43 |
| **Both tissues** | 1 | 0 | 7 | 4 | 8 | 7 |

### Meta-analysis of DGE in Blood

In blood, the DGE meta-analysis only reported the KIR2DL3 gene, which encodes for a transmembrane glycoprotein expressed by natural killer cells and subsets of T cells, as significantly altered in the SDID comparison. KIR2DL3 displays increased expression in MS females compared to MS males. In particular, the IDM comparison revealed lower KIR2DL3 expression in MS males than in healthy control males but did not show a significant change in the IDF comparison (Figure 3 and Figure 4, red box). This is a special case of Supplementary Figure 1D.

**Figure 4.**
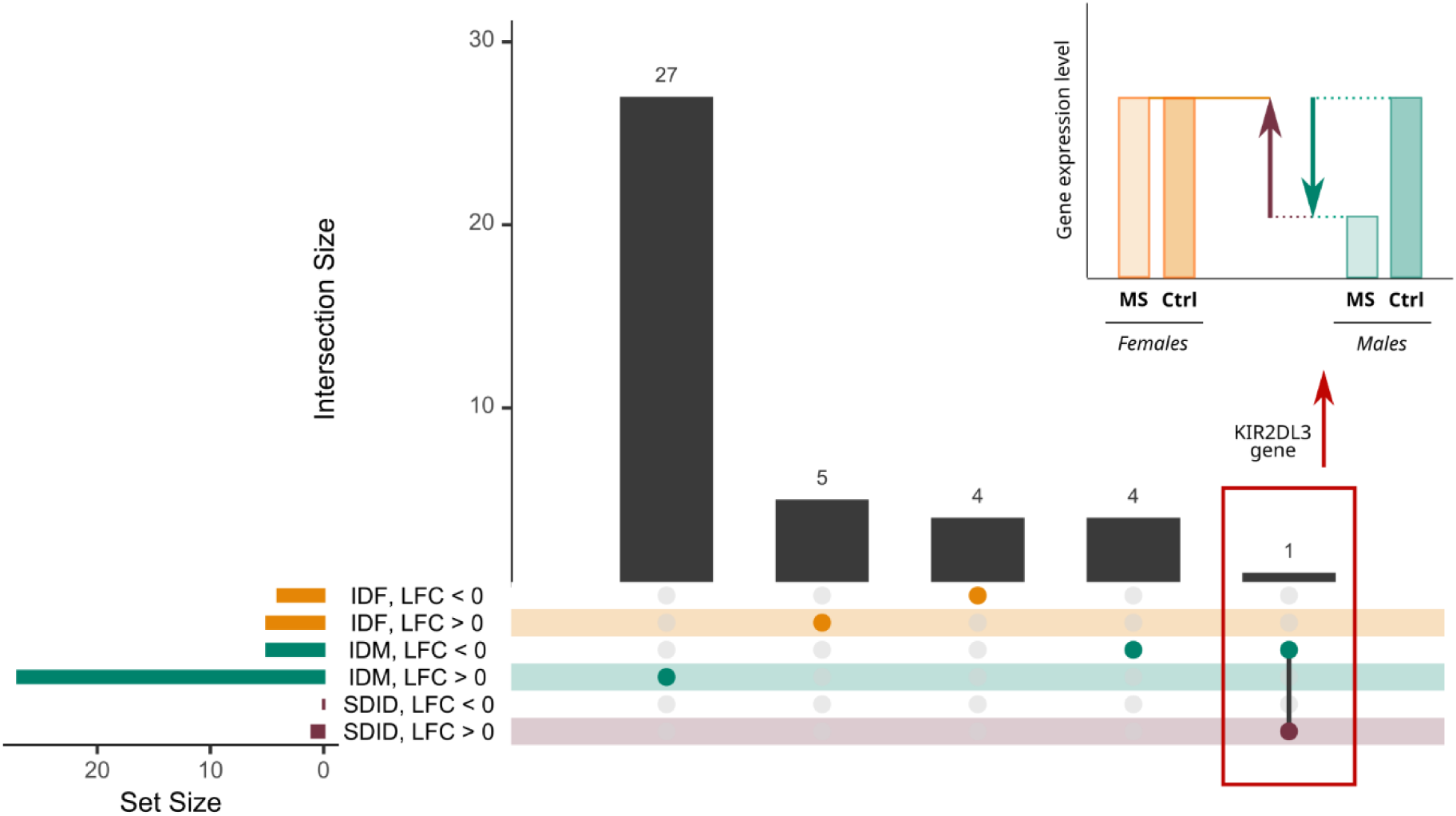
Upset plot of meta-analysis results in blood studies. The results of each comparison are separated according to the LFC sign. Horizontal bars indicate the number of significant genes in each comparison (1 specific color for each comparison). The vertical bars indicate the genes included in the intersection of the groups denoted with a colored dot underneath. A colored dot under a bar indicates that the genes are specific to this group. The red box indicates the case of the KIR2DL3 gene, which is significant SDID and IDM comparisons in the manner represented by the bar plot in the upper right corner.

The IDF and IDM comparisons reported 40 significant sex-specific genes not implicated in the SDID comparison (Figure 4 and Table 5). Interestingly, some genes have known associations with MS in the Open Targets Platform (Table 5, in bold), while other genes, such as PSMB2, SRP14, TESC, and KAT7, have known associations with neurodegenerative diseases, the innate immune system, neuroprotective effects, and T cell development, respectively [38–41].

**Table 5.**
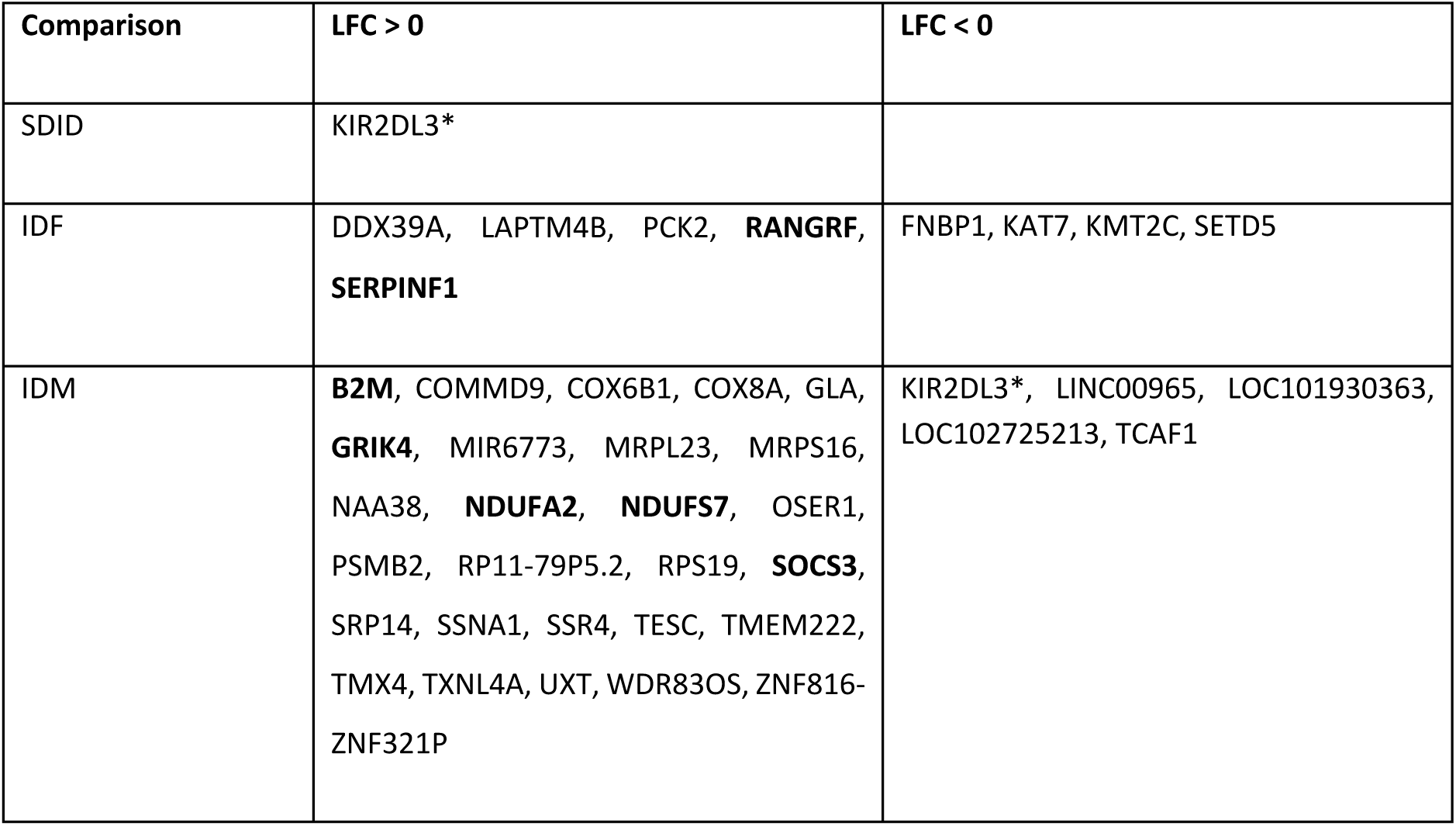
Meta-analysis results of blood samples. Significant genes for SDID (sex-differential impact of the disease), IDF (impact of the disease in females), and IDM (impact of the disease in males) comparisons. The results are separated into columns according to the LFC sign of the genes. Genes associated with MS in the Open Targets database are indicated in boldface. *KIR2DL3 was not associated with MS in Open Targets but was in a previous publication [42].

### Meta-analysis of DGE in Nervous Tissue

In nervous tissue, our meta-analysis detected 13 significant genes in the SDID comparison (Table 6). Of these 13, 6 genes (RNF10, IFFO2, SLC17A5, UBXN2B, ZNF117, and ZNF488) have previous links to neurodegenerative diseases, while IFFO2 participates in muscle control [43–48] and 5 other genes (CECR7, STMP1, TRAF31P2-AS1, and LOC401127) [49–52] possess links to inflammation, bone, and autoimmune diseases. The remaining two genes - CEP78 and NUDT18 - encode for a centrosomal protein and a hydrolase of the Nudix superfamily, respectively. Interestingly, the CECR7 gene displayed significance in all 3 comparisons (Figure 5, red box and upper right corner), being overexpressed in MS females and healthy males.

**Table 6.**
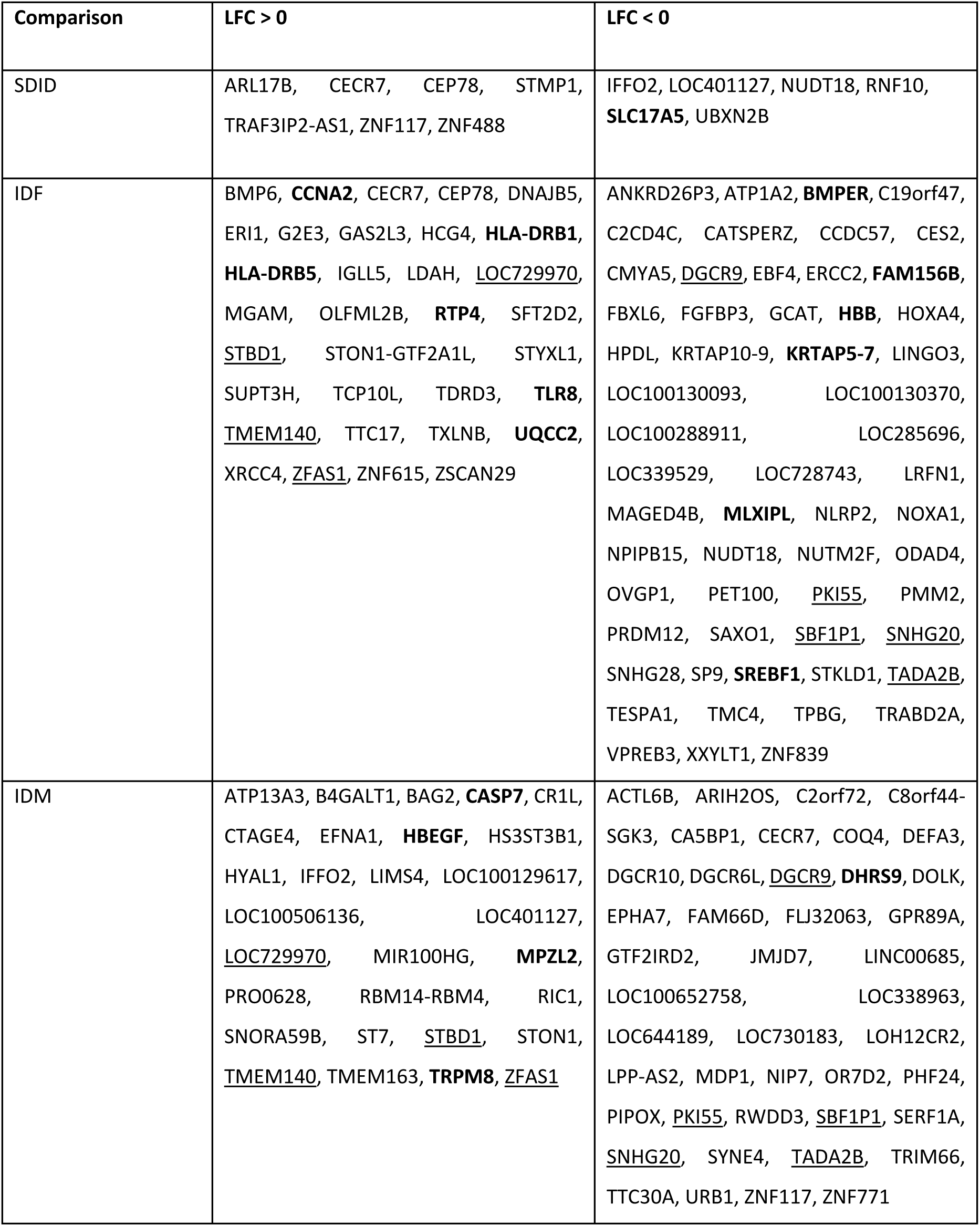
Meta-analysis results from nervous tissue samples. Significant genes for SDID (sex-differential impact of the disease), IDF (impact of the disease in females), and IDM (impact of the disease in males) comparisons. The results are separated into columns according to the LFC sign of the genes. Genes associated with MS in the Open Targets database are indicated in boldface. Genes altered in the same direction in the IDF and IDM comparisons are underlined.

**Figure 5.**
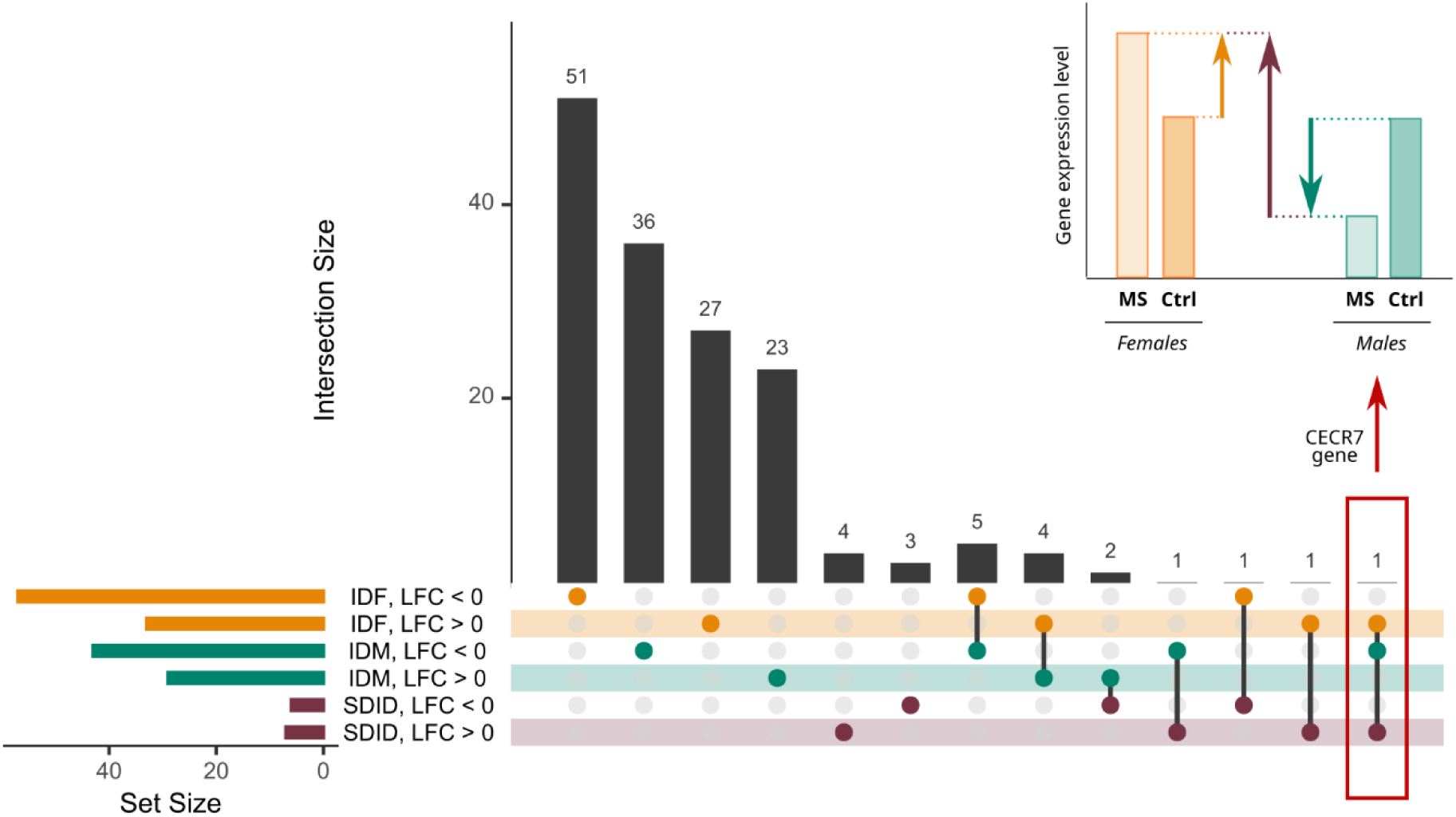
Upset plot of the meta-analysis results in nervous tissue samples. The results of each comparison are separated according to the LFC sign. Horizontal bars indicate the number of significant genes in each comparison (a specific color for each comparison). The vertical bars indicate the genes included in the intersection of the groups denoted with a colored dot underneath. A colored dot under a bar indicates that the genes are specific to this group. The red box indicates the case of the CECR7 gene, which is significant in all comparisons in the manner represented by the bar plot in the upper right corner.

We found 90 and 72 significant genes in the IDF and IDM comparisons, respectively (Figure 5). In the IDF and IDM comparisons, 9 genes displayed the same pattern of change, suggesting that they may represent possible sex-independent disease markers (Figure 5). Table 6 describes all significant genes in nervous tissue, indicating those associated with MS according to the Open Targets Platform (in bold) and those common to the IDF and IDM comparisons (underlined). We display a functional enrichment analysis in the following sections, given the high number of genes involved in IDF and IDM comparisons.

### Blood and Nervous Tissue Meta-analysis of DGE

The two-tissue meta-analysis revealed an altered gene in the SDID comparison (LOC102723701), whose expression increases in females (Table 7). This lncRNA class is described as an antisense transcript of ERLIN2, a gene associated with juvenile primary lateral sclerosis and immune cells infiltration [53–55]. We found 11 and 15 genes significantly altered in the IDF and IDM comparisons, respectively; however, none displayed commonality (Figure 6, Table 7). We highlight KAT7 (also significant in the blood meta-analysis), SERF2 (a positive regulator of myeloid protein aggregation) [56], and NTN3 (a netrin, which controls guidance of CNS commissural axons and peripheral motor axons) [57] as well as others previously associated with disease (highlighted in bold in Table 7).

**Table 7.** Results from the two-tissue meta-analysis. Significant genes for SDID (sex-differential impact of the disease), IDF (impact of the disease in females), and IDM (impact of the disease in males) comparisons. The results are separated into columns according to the LFC sign of the genes. Genes associated with MS in the Open Targets database are indicated in boldface.

| Comparison | LFC > 0 | LFC < 0 |
| --- | --- | --- |
| SDID | LOC102723701 |  |
| IDF | BANF1, **CCL19**, **GABRE**, LOC101927972, NTN3, RPL35, SERF2 | **EP400**, FNBP1, KAT7, ZZEF1 |
| IDM | B4GALT1, **DUSP1**, DUSP18, FAU, **RPL19**, TXNL1, VDAC2, ZNF844 | A2M-AS1, FLJ30403, LOC283861, LOC286272, LOC400968, LOC728903, OR7D2 |

**Figure 6.**
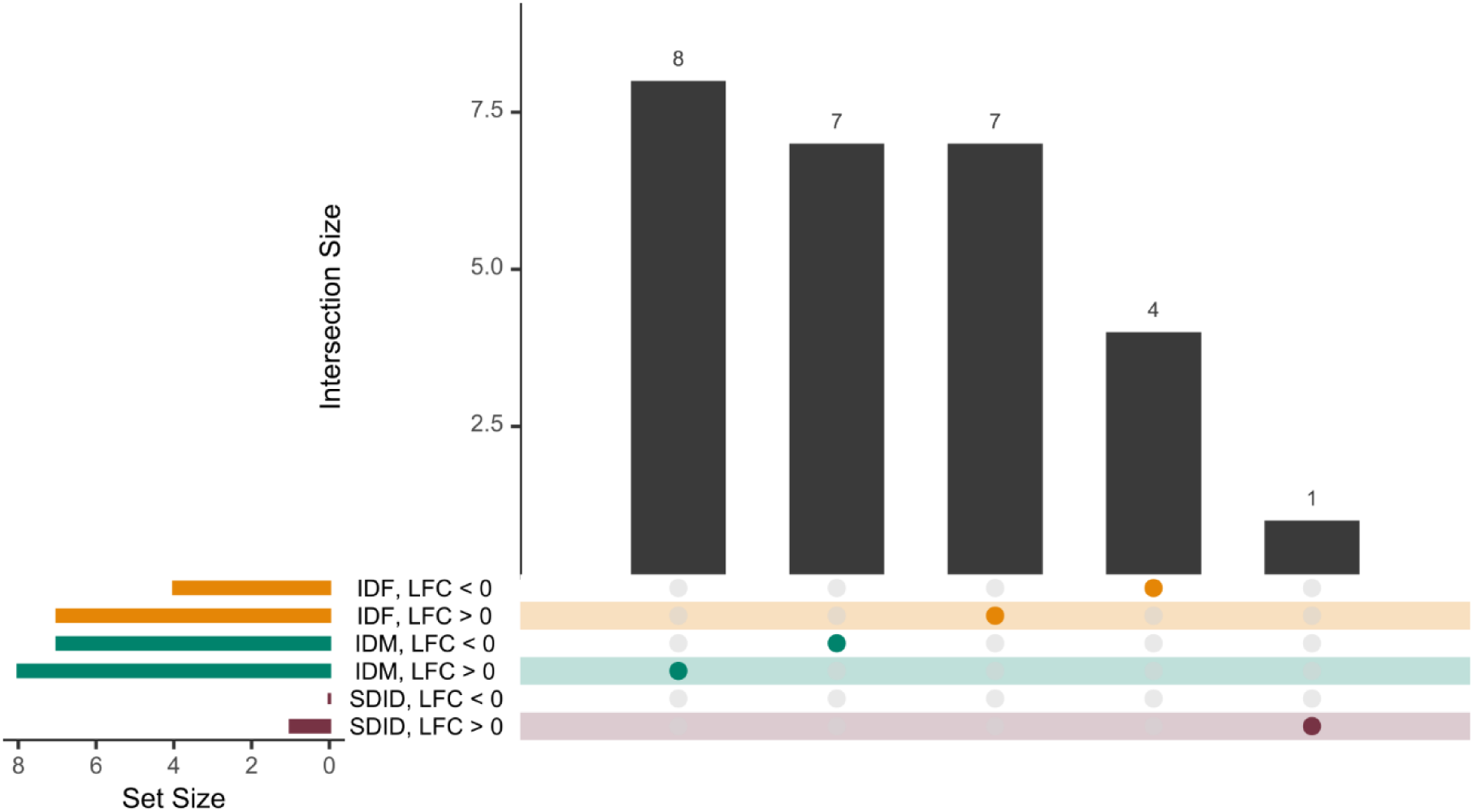
Upset plot of the two-tissue meta-analysis results. The results of each comparison are separated according to the LFC sign. Horizontal bars indicate the number of significant elements in each comparison (one specific color for each comparison). The vertical bars indicate the genes included in the intersection of the groups denoted with a colored dot underneath. A colored dot under a bar indicates that the genes are specific to this group.

### Functional Enrichment of DGE Meta-analysis in Nervous Tissue

We performed a GSA on the genes altered in all comparisons (IDF, IDM, and SDID) of the nervous tissue meta-analyses to provide a functional and integrative perspective of these genes in BP GO terms and KEGG pathways. In the SDID comparison, we detected the alteration of the “Staphylococcus aureus infection” KEGG pathway (hsa05150), which includes genes such as MHC class II antigen and Interleukin (IL-)10. In the IDF and IDM comparisons, we found significant results for both BP GO terms and KEGG pathways (Figure 7). We detected sex-specific elements and elements common to both sexes in both ontologies. Detailed results of the functional enrichment can be found in the supplementary material, the Zenodo repository, and the metafun-MS web platform.

**Figure 7.**
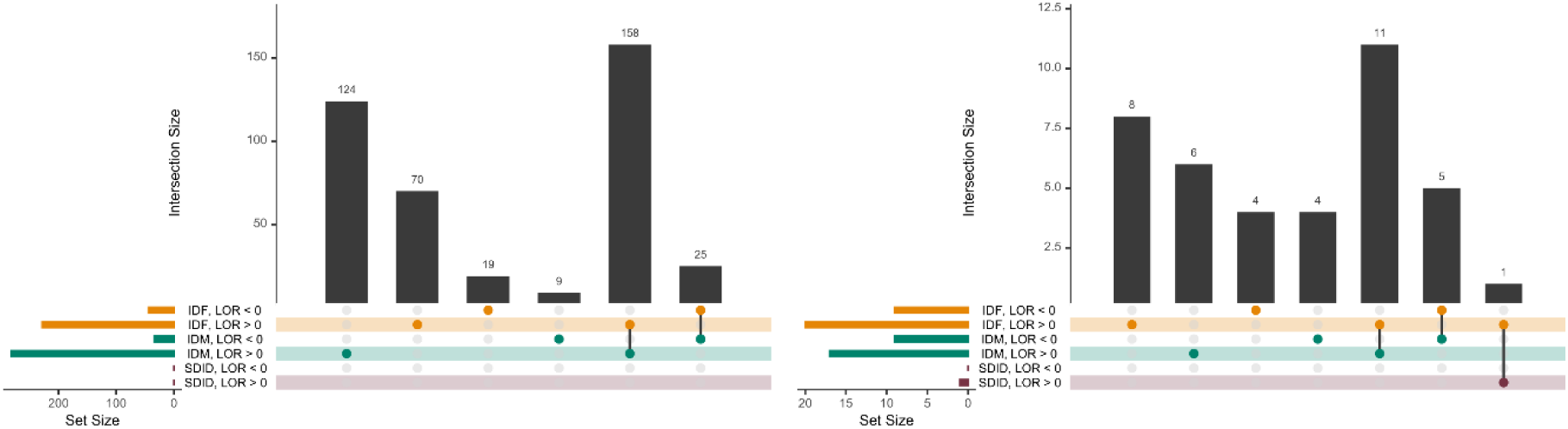
Upset plot of the functional enrichment results in nervous tissue for BP GO terms. Horizontal bars indicate the number of significant BP GO terms in each comparison (a specific color for each comparison). Results are exhibited separately for significantly upregulated (LOR > 0) and downregulated (LOR < 0) BP GO terms. The vertical bars indicate the genes included in the intersection of the groups denoted with a colored dot underneath. A colored dot under a bar indicates that the genes are specific to this group.

To summarize the high number of significant BP GO terms in the IDF and IDM comparisons, we classified them into 15 manually selected parental GO classes (hierarchically superior GO terms) (Figure 8). We classified BP GO terms with no ancestor among the selected parental GO classes as “Other.” Interestingly, the number of BP GO terms overrepresented in patients with MS (LOR > 0, Figure 8) is notably superior to those overrepresented in healthy patients (Figure 8, LOR < 0). The parental GO classes with the highest number of alterations were “Immune system process”, “Response to stimulus” and, to a lesser extent, “Cellular response to stimulus” and “Anatomical structure development”. Regarding the immune system, MS females specifically presented altered BP GO terms related to the myeloid lineage, regulation, and production of cytokines (IL-1, 2, 6, and 10), while MS males presented alterations in the lymphocyte lineage (Figure 9). MS females also presented altered mitochondrial respiratory chain complexes and purine and glutamate metabolism, while MS males presented alterations in amine and amino acid transport and stress response to metal ions (Figure 9).

**Figure 8.**
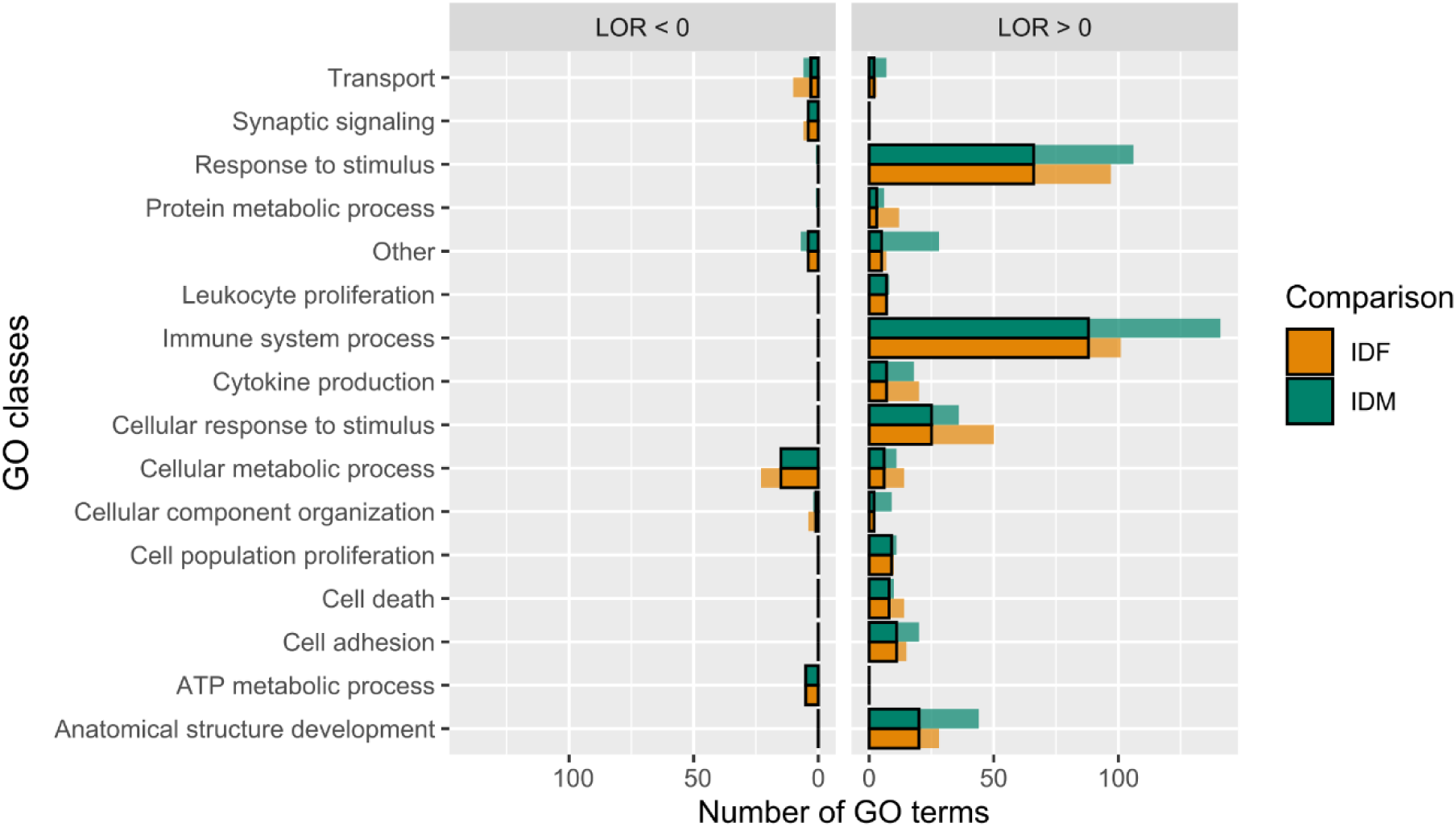
Bar chart of the number of significant BP GO terms in the IDF and IDM comparisons. The horizontal bars show the number of BP GO terms, separated between overexpressed in MS patients (LOR > 0, right) and healthy patients (LOR < 0, right) in each parental GO class. The orange and green bars (IDF and IDM, respectively) show the number of GO terms within each parental GO class. Opaque bars surrounded by a black border indicate items common to the comparisons, while translucent bars indicate items unique to IDF or IDM.

**Figure 9.**
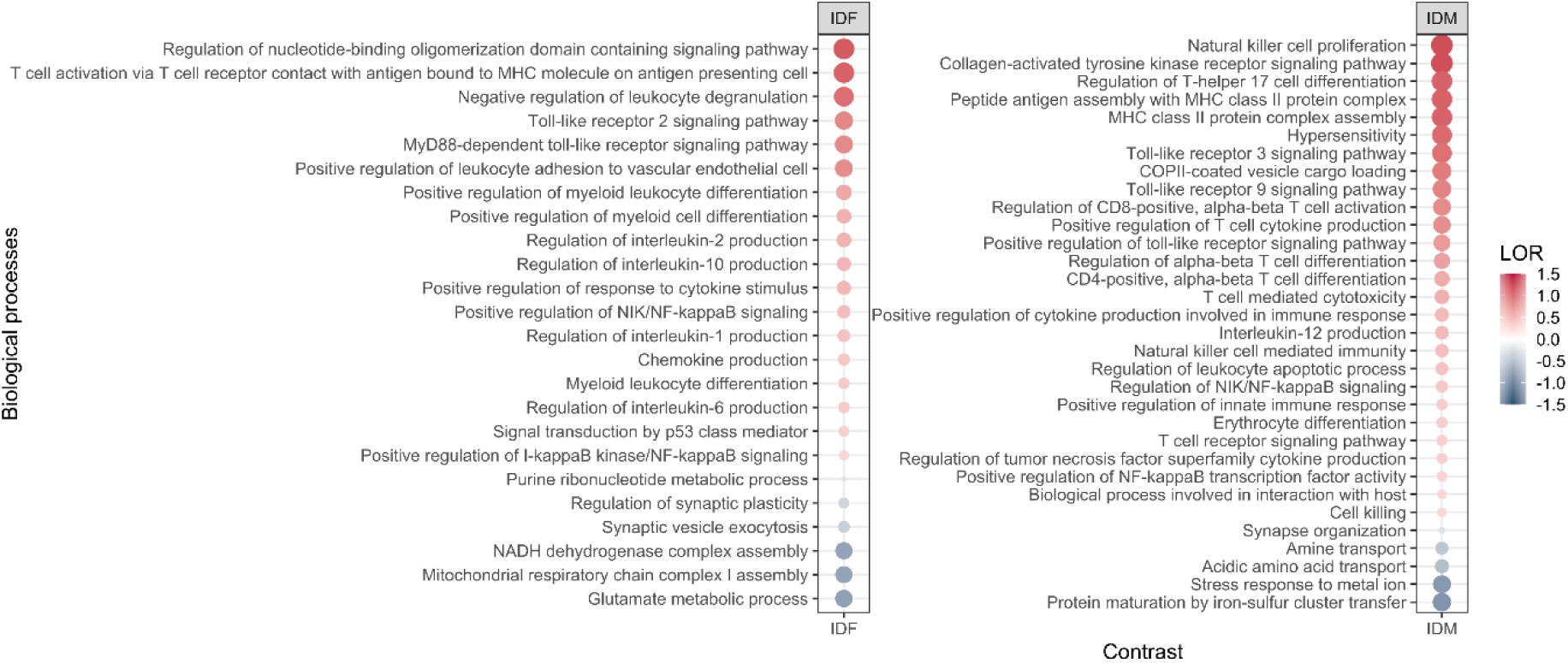
Dotplot of a representative selection of the specific significant BP GO terms in IDF and IDM. GO terms affected exclusively in IDF (left) or IDM (right). The color and size of the dots represent the sign and magnitude of the change (LOR).

In the case of IDF, the KEGG analysis demonstrated altered signaling pathways via Toll-like receptor and NOD-like receptor (constitute two major forms of innate immune sensors), as well as osteoclast differentiation, p53 pathway, and metabolism pathways related to biotin, alanine, aspartate and glutamate, xenobiotics, and drugs. KEGG pathway analysis of the IDF comparison also demonstrated alterations in terms related to diseases other than MS, including Chagas disease, prostate cancer, and Staphylococcus aureus infection. The IDM comparison showed alterations in KEGG pathways related to cytokine-cytokine receptor interactions, the Jak-STAT signaling pathway, protein processing in the endoplasmic reticulum, the peroxisome, olfactory transduction, calcium signaling, metabolism related to valine, leucine and isoleucine degradation, butanoate, and diseases such as small cell lung cancer and auto-immune thyroid disease (Supplementary Table 9).

### Metafun-MS platform

We have developed an open-source web platform, Metafun-MS (http://bioinfo.cipf.es/metafun-MS), containing a detailed description of the 9 datasets and 474 samples involved in this study. This resource includes detailed information on all genes, BP GO terms, and KEGG pathways that users can explore to identify transcriptomic profiles of interest for each study.

For each significant gene in a meta-analysis, Metafun-MS depicts the global activation level for all studies and each study’s specific contribution using statistical indicators (LFC/LOR, confidence interval, p-value, and adjusted p-value) and graphical representations by function (forest and funnel plots). This open resource aims to contribute to data sharing between researchers, the elaboration of innovative studies, and the discovery of new findings.

## Discussion

A range of previous studies have integrated data from numerous sources to characterize sex-based differences [58–60]; however, to our knowledge, this study represents the first meta-analysis of transcriptomic studies in MS and the first identification of sex-specific biomarkers in several tissues, which provides a better understanding of the molecular mechanisms underlying MS. This pathology has an important impact on society as one of the significant causes of disability in young adults; however, data regarding the causes of MS remain limited. Furthermore, the significant variability in clinical manifestations makes diagnosis and prognosis challenging tasks [2]. MS remains a clinical challenge, as current treatments focus on symptom amelioration and the slowing of the clinical course of the disease rather than patient recovery.

We carried out an *in-silico* approach using MS data related to sex from public repositories as a rapid and cost-effective means to predict and validate the effects of relevant biological variables on human health and guide further clinical research. A lack of standardization in the description, nomenclature, and format of public data represented significant challenges to our approach. Therefore, in addition to encouraging free access to research data (Open Science), we also wish to promote the FAIR principles (Findable, Accessible, Interoperable, Reusable) [61], which aim to standardize the deposition of data in repositories.

In this study, we performed a meta-analysis using a robust random-effects model, which allows for the integration of information from individual studies to obtain greater statistical power and precision and reveals findings that cannot be easily obtained through the intersection or addition of results in individual studies. Table 3 depicts the diverse nature of each study - study GSE108000 in nervous tissue provided many differentially expressed genes, while others failed to provide any. This variability highlights the need to employ strategies such as meta-analysis to integrate information from individual studies and obtain a transcriptional consensus profile as a final result. We applied the same bioinformatics strategy from normalized expression matrices to DGE results with updated gene names to ensure comparable results and reduce biases. The design and application of this strategy identified sex-specific MS biomarkers. The further development of studies with sex information within each MS subgroup may extend the identification of biomarkers into specific subtypes and affected tissues.

### Meta-analysis of transcriptomics profiles

Since MS is a chronic autoinflammatory disorder of the CNS, we performed separate meta-analyses in blood and nervous tissue and a third meta-analysis that integrated both tissues. Identifying sex-based differences represented our primary goal by studying genes differentially altered in the SDID comparison. Meanwhile, the IDF and IDM comparison allowed us to detect specific effects in each sex, understand the basis of the disease for each sex, and find common patterns in both sexes.

### Meta-analysis in blood

Our meta-analysis in blood identified a single gene, KIR2DL3, in the SDID comparison. Interestingly, we also observed alterations to KIR2DL3 in the IDM comparison, with this gene downregulated in male MS patients. KIR2DL3 is related to MHC I mediated antigen processing and presentation and innate immune system pathways and has a protective effect; however, the absence of KIR2DL3 has been linked to MS development [42, 62]. Moreover, the protective role of KIR2DL3 has been documented in other autoimmune diseases such as rheumatoid vasculitis [63]. Identifying biomarkers in blood that facilitate the non-invasive diagnosis, prognosis, and evaluation of treatment response remains crucial for MS patients who occasionally undergo cerebrospinal fluid analysis. Unfortunately, identifying blood-based biomarkers remains challenging due to factors such as molecule availability in the blood, the dilution factor, sensitivity, specificity, and disease stage. Together, these factors may explain why we only identified a single gene, KIR2DL3, in the SDID meta-analysis in blood.

The blood meta-analysis in IDF comparison revealed a gene dysregulation profile related to the immune system and symptoms associated with MS. For example, DDX39A, LAPTM4B, PCK2, RANGRF, SERPINF1, FNBP1, KAT7, KMT2C, and SETD5 displayed dysregulation in MS females. DDX39A and LAPTM4B may play roles in controlling immune responses [64, 65], while RANGRF has been related to cardiac arrhythmia in Brugada Syndrome [66]. The number of cardiac ventricular arrhythmias worsens the prognosis of MS patients with a higher risk of sudden death [67]. SERPINF1 may influence immune and vascular injury [68]. Studies have generally described roles in inflammation, immunity, and dementia for serpins, while serpin family members have been identified in MS plaques and neurodegenerative conditions such as Alzheimer’s disease [69]. FNBP1 may represent a significant biomarker, as the expression of this gene links to prognosis and level of immune infiltration in different cancers [70] and could play a similar role in MS. Interestingly, KAT7, which gene encodes a histone acetyltransferase crucial for the development, maintenance, and survival of T cells and the prevention of auto-immune diseases [71], appeared downregulated in MS females. This study represents the first description of a sex-based association of KAT7 with MS to the best of our understanding.

The IDM blood meta-analysis reflected the dysregulation of genes related to inflammation, immune system, and autoimmune diseases (B2M, COMMD9, GLA, PSMB2, SOCS3, SRP14, SSNA1, SSR4, TESC, UXT, and RPS19 upregulated in MS males and KIR2DL3 downregulated in MS males), genes related to redox activity/stress oxidative and mitochondrial function (COX6B1, COX8A, MRPL23, MRPS16, NDUFA2, NDUFS7, OSER1, TMX4, and TXNL4A upregulated in MS males) and genes related to neurodegenerative diseases and intellectual disabilities (WDR83OS, GRIK4, NAA38, TMEM222 upregulated in MS males). Other short and long non-coding (lnc)RNA genes (MIR6773, LINC00965, LOC101930363, and LOC102725213) could be critical for post-transcriptional regulation of other genes, while TCAF1, a transient receptor potential channel-associated factor, can regulate cell migration [72]. These results suggest that MS hallmarks such as inflammation and immunity occur in both MS females and males through different target genes in each case, which may be critical to guide medical decisions. Unfortunately, the blood meta-analyses failed to identify common MS biomarkers for males and females, perhaps due to difficulties finding stable markers in the blood, as discussed above.

### Meta-analysis in nervous tissue

Analyses in nervous tissue yielded a higher number of differentially expressed genes than in blood, reflecting the notable impact of MS in this tissue. The SDID nervous tissue meta-analysis found 13 differentially affected genes by sex, with specific genes related to the loss of mental functions and neural disorders, blood cell abnormalities, inflammation, and autoimmune disorders.

ARL17B, CECR7, CEP78, STMP1, TRAF31P2-AS1, ZNF117, and ZNF488 displayed increased expression in MS females, as did the CECR7 and CEP78 genes (LFC > 0 in IDF). CECR7 has been previously related to immune infiltration in hepatocellular carcinoma [73] and regulates the expression of CTLA4 by targeting miR-429 [74]; meanwhile, CEP78 has been recognized as an auto-antigen in prostate cancer [75]. Thus, both genes represent potential candidates for immunotherapies. STMP1 has a reported involvement in the NLRP3 inflammasome and Paget’s disease bone [50], while TRAF31P2-AS1 participates in IL-17 regulation. Interestingly, this interleukin possesses pro-inflammatory properties and contributes to immune dysregulation in other autoimmune diseases such as colitis and rheumatoid arthritis [76].

IFFO2, LOC401127, NUDT18, RNF10, SLC17A5, or UBXN2B, presented a negative LFC in this SDID comparison. In particular, IFFO2 and LOC401127 displayed significant upregulation in MS males (LFC > 0 in IDM) while NUTD18 displayed a significant downregulation in MS females (LFC < 0 in IDF). The presence of intermediate filament-associated proteins (IFAPs) in MS plaques tissue has been previously documented [77]; therefore, IFFO2 may represent a novel IFAP for MS male plaque tissues. The LOC401127 lncRNA could play an essential role in MS regulation or prognosis, as for other described lncRNAs in chronic musculoskeletal disorders [78]. We also highlight the protective role against oxidative stress and DNA damage of NUTD18 (also known as MTH3), which displays downregulation in MS females [79].

In the IDF and IDM meta-analyses in nervous tissue, we identified genes that appeared dysregulated in both sexes; therefore, these genes may represent potential MS biomarkers - LOC729970, STBD1, TMEM140, and ZAFS1 displayed upregulation and DGCR9, PKI55, SBF1P1, SNHG20, and TADA2B displayed downregulation in MS in both sexes. Specific genes identified in the IDF and IDM comparisons have not been previously linked to MS and may represent interesting future research directions. In general, we observed an increase in genes related to inflammatory and immune responses in female MS patients. Autoimmune diseases are generally exacerbated in female patients, and a more pro-inflammatory phenotype of XX mice than XY mice has been demonstrated in autoimmune diseases such as experimental and spontaneous lupus models [80, 81]. Sex-based differences in immune responses are well-documented and influenced by multiple factors (chromosomes organization, reproductive organs, and sex hormone levels), which, in turn, influence the incidence and other aspects of autoimmune diseases [82].

**The two-tissue meta-analysis** identified a single gene - LOC102723701 - as displaying SDID. This lncRNA gene, not previously related to MS, is described as a novel transcript antisense to the ERLIN2 gene. The misfunction of ERLIN2, which mediates intracellular calcium signaling and participates in neurodevelopment and neurotransmission, leads to motor disorders and spastic paraplegia [83, 84]. Importantly, ERLIN2 mutations lead to abnormal splicing and nonsense-mediated mRNA decay, promoting motor neurodegeneration and juvenile amyotrophic lateral sclerosis [55]. Additionally, ERLIN2 has been related to the infiltration of immune cells in the tumor microenvironment [54]. Therefore, LOC102723701 may represent an exciting and novel target in MS for future research. Regarding the IDF and IDM comparisons in both tissues, some of the identified genes (e.g., CCL19, GABRE, and EP400 in IDF; DUSP1 and RPL19 in IDM) have been previously associated with MS in the Open Targets platform, thereby supporting our results. In general, the two-tissue meta-analysis mirrored the results from the meta-analysis in individual tissues. Genes related to oxidative stress, mitochondrial functions, metabolism, immune system, and other neural or autoimmune disorders such as Alzheimer’s disease or lupus erythematosus displayed dysregulation in MS patients.

Focusing on the **functional enrichment in nervous tissue**, we found a single significantly dysregulated KEGG pathway in the SDID comparison: “Staphylococcus aureus infection” that involved important immune genes important immune genes (MHC class II antigen and IL-10). We also discovered the upregulation of this pathway in MS females, further supporting the differential role of the immune system in MS between males and females. When we analyzed GO terms in the IDF and IDM comparisons, many biological processes related to inflammation and immunity displayed sex-dependent dysregulation in MS patients. Specifically, MS females presented alterations related to myeloid linage and pro-inflammatory environment, influenced by alterations in interleukin regulation and response (e.g.., IL-1, IL-2, IL-6, IL-10, and interferon-gamma). These alterations are closely related to innate immune responses and have been previously linked to MS and autoimmune diseases [85–89]. In contrast, MS males presented immune alterations related to immunomodulation of lymphoid linage (e.g., T cell subsets, natural killer cells, dendritic cells), affecting activation, proliferation, or differentiation, which have a closer relation to adaptative immune responses [90–92]. Identifying T cell clones associated with human autoimmunity remains challenging and could have critical connotations in MS risk and outcomes. Taken together, the different scenarios observed in the immune system could partly explain sex-based differences in susceptibility to MS. Pro-inflammatory cytokines, which are significantly altered in MS females, have been related to autoimmune disorders such as arthritis rheumatoid [93]. Meanwhile, MS males suffer from a more rapid and worsened progression, which could be explained by more intensive participation of T cells in demyelination. Further studies must be carried out to clarify these hypotheses and help guide decisions in the design of personalized immunotherapies.

Other sex-related differences in GO terms included alterations in mitochondrial respiratory chain complexes, purine, and glutamate metabolism in MS females and alterations in amine and amino acid transport and stress response to metal ions in MS males. These biologic processes have important connotations in MS - the glutamatergic system plays a vital role in neurotoxicity and MS pathogenesis [94], while amino acid metabolism affects immune homeostasis, as activated immune cells present a high demand for amino acids [96].

Analysis of the KEGG pathway data revealed that MS females displayed an upregulation in several pathways related to immune signaling receptors (Toll-like receptor signaling pathway, NOD-like receptor signaling pathway, B cell receptor signaling pathway), the upregulated pro-inflammatory microenvironment observed in GO terms, and the innate responses explained above. Toll-like and Nod-like receptors are two major forms of innate immune sensors and provide immediate responses against pathogenic invasion or tissue injury [99, 100]. Activation of these sensors induces the recruitment of innate immune cells such as macrophages and neutrophils (myeloid linage) (which displayed dysregulation in MS females), initiates tissue repair processes, and results in adaptive immune activation. Abnormalities in these innate sensor-mediated processes may cause excessive inflammation due to hyper-responsive innate immune signaling or sustained compensatory adaptive immune activation [101]. Other KEGG pathways enriched in MS females have associations with chronic or autoimmune diseases (e.g., chronic myeloid leukemia, systemic lupus erythematosus, and rheumatoid arthritis), which may indicate that autoimmune diseases share altered signaling pathways. We also discovered the upregulation of intestinal immune networks for IgA production and Chagas disease in MS females. Diet, microbiota gut, parasite, and viral infections are associated with chronic inflammatory diseases and could play a significant role in MS etiology [102]; meanwhile, intestinal disorders also represent common symptoms of MS [103, 104]. The KEGG pathways downregulated in MS females include the metabolism of xenobiotics by cytochrome P450, drug metabolism - cytochrome, cardiac muscle, and Alzheimer’s disease, which could have significant implications in treatment responses and therapy development. In MS males, upregulated KEGG pathways related to RNA transport, cytokine-cytokine receptor interaction, protein processing in the endoplasmic reticulum, the phagosome, JAK-STAT signaling pathway, and olfactory transduction, while valine, leucine, and isoleucine degradation represent the unique downregulated pathway. RNA localization controls gene expression, while neurological diseases such as amyotrophic lateral sclerosis and fragile X syndrome have been associated with dysregulated RNA localization in neurons [105]. Endoplasmic reticulum-associated protein modifications, folding, and degradation could mediate the pathogenesis of autoimmune disorders [106], while antigen processing and cross-presentation by dendritic cells require phagosome maturation [107]. Finally, metabolism and diet have relevant implications in MS, as in other autoimmune diseases [108], so amino acid degradation may impact MS at this level.

### Strengths and limitations

Patient sex influences the observed pathophysiological differences in neurodegenerative and autoimmune diseases [3, 109], with sex-based differences described in MS risk and disease progression [110]. Unfortunately, sex is not frequently considered a biological variable during biomedical research; this became evident during our systematic review, where the absence of sex information prompted us to discard a substantial number of studies. The small number of studies with sex information has not allowed us to stratify MS subtypes; however, our global approach could identify a set of biomarkers common to MS. Encouragingly, the sex perspective has gained recent relevance as a crucial aspect of personalized medicine and has been recognized as a necessary biological variable for inclusion in research by the National Institutes of Health (US Department of Health and Human Services) [111] and the European Union [112, 113].

## Conclusions

We have performed a gene and functional meta-analysis of 9 MS transcriptomic studies from a sex perspective, identifying sex-based differences and similarities at gene, function, and pathway levels. The obtained results allow for a better understanding of those mechanisms crucially involved in MS and the identification of potential biomarkers, whose detailed study may improve personalized therapies. In addition, we highlight the importance of providing information on sex in studies and the need for open platforms for data sharing to allow for efficient advancements in research.

## Data Availability

The large volume of data and results generated in this study are freely available through the Metafun-MS web tool: http://bioinfo.cipf.es/metafun-MS and the zenodo repository: http://doi.org/10.5281/zenodo.6344450. This study analyzed transcriptomic data available in the Gene Expression Omnibus database with accession numbers GSE37750, GSE41848, GSE41849, GSE41890, GSE62584, GSE108000, GSE111972, GSE131281, and GSE135511.

## Additional files

Detailed results of all analyses and the versions of the R packages used can be found in the Zenodo repository: http://doi.org/10.5281/zenodo.6344450

## Abbreviations

BH: Benjamini & Hochberg p-value adjust method
BP: Biological processes of Gene Ontology
CIS: Clinical Isolated Syndrome
CNS: central nervous system
DGE: Differential gene expression
GEO: Genes Omnibus Expression database
GO: Gene Ontology
GSA: Gene set analysis
IDF: Impact of the disease in females
IDM: Impact of the disease in males
IL: Interleukin
KEGG: Kyoto Encyclopedia of Genes and Genomes
LFC: Logarithm of the fold change
LOR: Logarithm of odds ratio
MS: Multiple sclerosis
PCA: Principal component analysis
PRISMA: Preferred Reporting Items for Systematic Reviews and Meta-Analyses
PPMS: Progressive Primary Multiple Sclerosis
PRMS: Progressive-Relapse Multiple Sclerosis
RRMS: Relapsing-Remitting Multiple Sclerosis
SDID: Sex-differential impact of the disease
SPMS: Secondary-Progressive Multiple Sclerosis

## Availability of data and materials

The large volumes of data and results generated in this study are freely available through the Metafun-MS web tool: http://bioinfo.cipf.es/metafun-MS and the Zenodo repository: http://doi.org/10.5281/zenodo.6344450. This study analyzed transcriptomic data available in the Gene Expression Omnibus database with accession numbers GSE37750, GSE41848, GSE41849, GSE41890, GSE62584, GSE108000, GSE111972, GSE131281, and GSE135511.

## Computer Code and Software

The code developed for the analyses described in this work and the software and its versions are publicly available at Zenodo: http://doi.org/10.5281/zenodo.6344450.

## Authors’ contributions

Conceptualization, F.G.-G.; Data curation, J.F.C.-S., F.J.R., A.N.-A., and I.S.-S.; Formal analysis, J.F.C.-S., and F.J.R.; Funding acquisition, F.G.-G; Investigation, J.F.C.-S., Z.A., M.R.H., N.Y.-C., and F.G.-G.; Methodology, J.F.C.-S., and F.G.-G.; Project administration, F.G.-G.; Software, J.F.C.-S., and M.R.H.; Supervision, F.G.-G.; Validation, Z.A., and N.Y.-C.; Visualization, J.F.C.-S., I.S.-S., M.R.H., and F.G.-G.; Writing – original draft, J.F.C.-S., Z.A., and F.G.-G.; Writing – review & editing, J.F.C.-S., Z.A., F.J.R., M.R.H., A.N.-A., I.S.-S., A.L.-C., N.Y.-C., M.I.-V., and F.G.-G. All authors have read and agreed to the published version of the manuscript.

## Acknowledgments

The authors thank the Principe Felipe Research Center (CIPF) for providing access to the cluster, co-funded by European Regional Development Funds (FEDER) in Valencian Community 2014-2020. The authors also thank Stuart P. Atkinson for reviewing the manuscript.

## Rights and permissions

Open Access This article is licensed under a Creative Commons Attribution 4.0 International License, which permits use, sharing, adaptation, distribution, and reproduction in any medium or format, as long as you give appropriate credit to the original author(s) and the source, provide a link to the Creative Commons license, and indicate if changes were made. The images or other third-party material in this article are included in the article’s Creative Commons license unless indicated otherwise in a credit line to the material. If material is not included in the article’s Creative Commons license and your intended use is not permitted by statutory regulation or exceeds the permitted use, you will need to obtain permission directly from the copyright holder. To view a copy of this license, visit http://creativecommons.org/licenses/by/4.0/.

