## Supplementary figures and images for "A Deep Transcriptome Meta-Analysis Reveals Sex-based Molecular Differences in Multiple Sclerosis"

### SupFig01_Comparisons_arrows.png

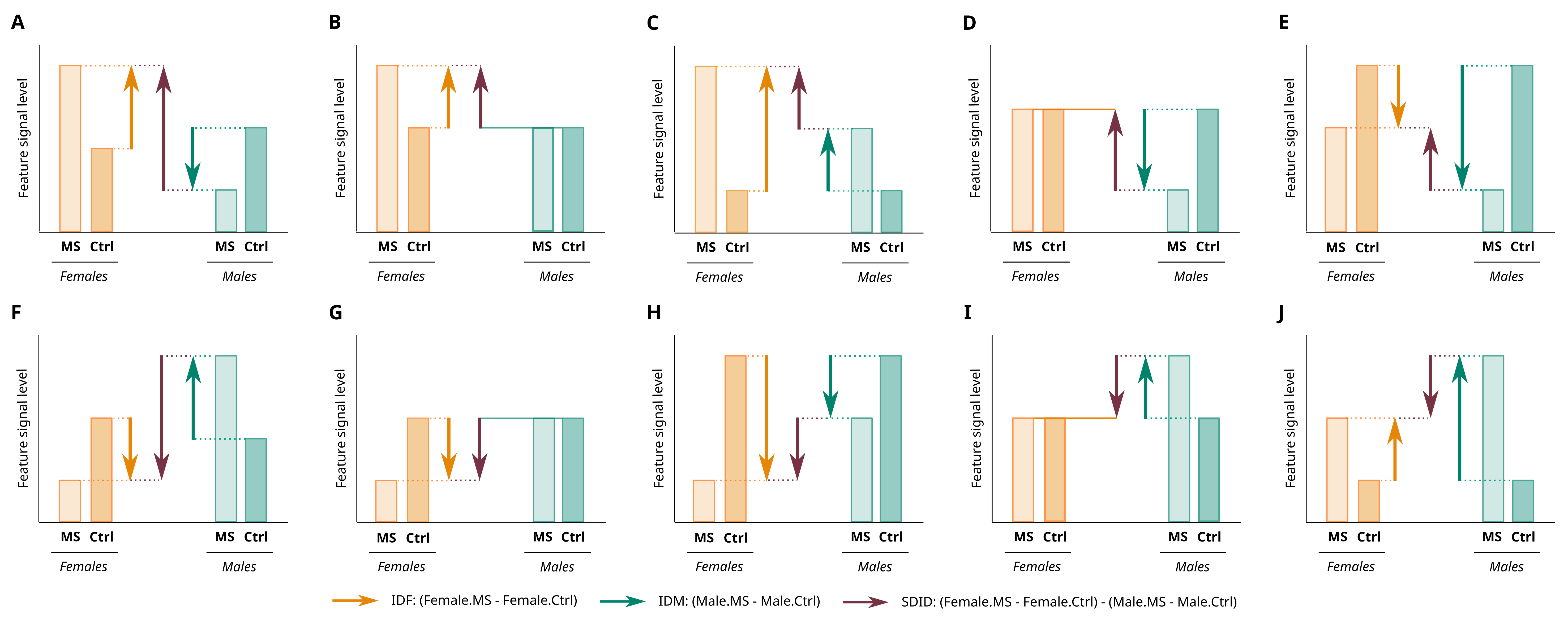
